# Assessing the causal effect of air pollution on risk of SARS-CoV-2 infection

**DOI:** 10.1101/2023.10.26.23297598

**Authors:** Annalan M D Navaratnam, Sarah Beale, Yamina Boukari, Vincent Nguyen, Wing Lam Erica Fong, Isobel Braithwaite, Thomas E Byrne, Ellen Fragaszy, Cyril Geismar, Jana Kovar, Parth Patel, Madhumita Shrotri, Alexei Yavlinsky, Andrew C Hayward, Haneen Khreis, Robert W Aldridge

**Author notes:** Correspondence to: Dr Annalan Navaratnam.

## Abstract

**Introduction:** Emerging evidence suggests association of air pollution exposure with risk of SARS-CoV-2 infection, but many of these findings are limited by study design, lack of individual-level covariate data or are specific to certain subpopulations. We aim to evaluate causal effects of air pollution on risk of infection, whilst overcoming these limitations.

**Methods:** Concentrations for black carbon(BC), particulate matter 10(PM_10_), particulate matter 2.5(PM_2.5_), nitrogen dioxide(NO_2_) and oxides of nitrogen(NO_x_) from the Department of Environment, Food and Rural Affairs (DEFRA) and Effect of Low-level Air Pollution: A Study in Europe (ELAPSE) were linked to postcodes of 53,683 Virus Watch study participants. The primary outcome was first SARS-CoV-2 infection, between 1st September 2020 and 30th April 2021. Regression analysis used modified Poisson with robust estimates, clustered by household, adjusting for individual (e.g., age, sex, ethnicity) and environmental covariates(e.g., population density, region) to estimate total and direct effects.

**Results:** Single pollutant analysis showed the direct effect of higher risk of SARS-CoV-2 infection with increased exposure to PM_2.5_(RR1.11,95%CI 1.08;1.15), PM_10_(RR1.06,95%CI 1.04;1.09), NO_2_(RR1.04,95%CI 1.04;1.05) and NO_x_(RR1.02,95%CI 1.02;1.02) per 1µg/m^3^ increment with DEFRA 2015-19 data. Sensitivity analyses altering covariates, exposure window and modelled air pollution data source produced similar estimates. Higher risk of SARS-CoV-2 per 10^-5^m^-1^ increment of BC (RR1.86, 95%CI 1.62;2.14) was observed using ELAPSE data.

**Conclusion:** Long term exposure to higher concentrations of air pollutions increases the risk of SARS-CoV-2 infection, highlighting that adverse health effects of air pollution is not only limited to non-communicable diseases.

## Introduction

Air pollution has become recognised as a major environmental threat to human health, not only representing a public health crisis, but highlights an inequity both in the UK and internationally.^1^ It is an established carcinogenic, including non-respiratory cancers, and there is increasing evidence of systemic inflammation and oxidative stress as important pathways for cardiovascular and metabolic disease. ^2–4^ Since the emergence of a global pandemic caused by SARS-CoV-2, resources and funding have been funnelled into finding explanations for geographical and demographic heterogeneity in COVID-19 incidence and deaths.^5^ Emerging epidemiological findings suggest involvement of air pollution in risk and severity of SARS-CoV-2 infection.^6^

There are three proposed mechanisms as to how long-term air pollution exposure is associated with higher risk of SARS-CoV-2 infection. Firstly, chronic exposure to air pollution is associated with higher risk of underlying respiratory, cardiovascular and metabolic conditions, which increase the risk of SARS-CoV-2 infection and worse clinical severity. Secondly, exposure to particulate matter (PM), nitrogen dioxide (NO_2_) and ozone (O_3_) may alter protein expression and alter host immunity to respiratory infections.^7,8^ PM exposure upregulates the expression of Angiotensin-Converting Enzyme 2 (ACE2), which the SARS-CoV-2 spike protein uses to bind to and enter host cells.^6,9,10^ NO_2_ exposure can lead to inflammation, increase cell permeability and impair tissue defences and phagocytic activity by depleting the antioxidant pool. ^11^ Finally, increased concentrations of PM_2.5_ (i.e., particulate matter less than 2.5micrometers in diameter) may increase the rate of COVID-19 transmission by facilitating viral transport over larger distances, although there is limited evidence for this mechanism.^12,13^

Much of the research into the association of air pollution and COVID-19 risk has been grounded in ecological studies, which compare population level results that may have systematic differences in testing and recording as well as lack available data on confounding factors.^14^ More evidence based on longitudinal studies with individual-level data and high resolution air pollution maps are needed to better understand this association.^14^ Kogevinas et al. (2021), using individual level data in a cohort study in Spain, found an association with risk of SARs-CoV-2 infection, but used symptom profiles in their criteria for case definition.^15^ Mendy et al (2021) used individual data from University of Cincinnati hospitals and clinics and demonstrated a 62% higher risk of hospitalisation in COVID patients per 1 µg/m^3^ increment in 10-year mean PM_2.5_ concentration.^16^ However, this was only in high risk patients, such as those with pre-existing asthma or chronic obstructive pulmonary disease.^16^ After adjusting for age, sex, socioeconomic status and comorbidities of every COVID-19 case and mortality in California, English et al identified those in the highest quintile of long-term PM_2.5_ exposure were at 20% and 51% higher risk of infection and mortality, respectively, compared to those in the lowest quintile.^17^ Covariates such as socioeconomic status and comorbidities were at the neighbourhood level.^17^

The Virus Watch study is a community cohort study across England and Wales, including serologically confirmed SARS-CoV-2 cases, which indicate prior SARS-CoV-2 infection regardless of symptom status at the time of infection. Virus Watch is not limited to clinical settings or specific populations and has individual level data on socioeconomic indicators and co-morbidities. This individual-level data, coupled with high-resolution air pollution maps, could provide further insight beyond association between air pollution and COVID-19 transmission. We aimed to estimate the causal effect of long term exposure to black carbon (BC), PM_10_, PM_2.5_, NO_2_ and oxides of nitrogen (NO_x_) on risk of SARS-CoV-2 infection, using adjustments for estimating total and direct effect.

## Methods

### Settings

The Virus Watch study is a household community cohort of acute respiratory infections in England and Wales that started recruitment in June 2020.^18^ As of 28th July 2022, 58,628 participants were recruited using a range of methods including post, social media and SMS messages and letters from their General Practice. Households were recruited from 24th June 2020 to March 2022 and asked to complete a post enrolment baseline survey containing demographic, medical history, financial and occupation questions. Individuals received a weekly illness survey via email to collect information on self-reported acute symptoms, vaccination status and PCR or lateral flow test results. Households also received a monthly survey of demographic, health-related, environmental and behavioural/psychosocial questions. Within the larger study, a sub cohort of 15,534 adults received monthly antibody testing.

### Outcome

The outcome was the first SARS-CoV-2 infection during the second wave in the UK, 1st September 2020 to 30th April 2021, during which the dominant variant in the UK was B.1.1.7 (i.e., alpha).^19,20^ This time period was selected as testing was not widely available during the first wave and vaccination was not widely available during the second wave, therefore minimising confounding factors that would need to be considered in the analysis. A SARS-CoV-2 case was identified based on the first positive result from the following:

1. Data linked to the Second Generation Surveillance System (SGSS), which contains SARS-CoV-2 test results using data from hospitalisations (Pillar 1) and community testing (Pillar 2). Linkage was conducted by NHS Digital using name, date of birth, address and NHS numbers, and sent in March 2021.The linkage period for SGSS Pillar 1 encompassed data from March 2020 until August 2021 and from June 2020 until November 2021 for Pillar 2.
2. Self-reported positive polymerase chain reaction (PCR) or lateral flow device (LFD) swabs for SARS-CoV-2 infection as part of the Virus Watch weekly survey.
3. Monthly self-collected capillary blood samples (400-600µL) in a subsample of 11,701 participants, which were tested in United Kingdom Accreditation Service (UKAS)-accredited laboratories. Serological testing using Roche’s Elecsys Anti-SARS-CoV-2 electrochemiluminescence assays targeting total immunoglobulin (Ig) to the Nucleocapsid (N) protein, or to the receptor binding domain in the S1 subunit of the Spike protein (S) (Roche Diagnostics, Basel, Switzerland). At the manufacturer-recommended seropositivity thresholds (≥1.0 cut-off index [COI] for N and ≥0.8 units per millilitre [U/ml] for S). A positive result was defined based on positivity to the N protein.
4. Clinical-collected venous blood samples tested for the S protein. In-clinic serology was conducted twice per participant between September 2020-January 2021 (Autumn round n = 3050) and April 2021-July 2021 (Spring round n = 2775)) (see study protocol for details).^18^ Positivity was defined as evidence of S-positivity in absence of receiving any COVID-19 vaccination prior to the serological test.

We used sliding date window matching (14 day window) to identify positive tests recorded by both Virus Watch and linkage to UK national records; where both were available, the linkage date was used. Where both swab and serological positives were recorded, we used the PCR/LFT date, unless the serological positive occurred first. Reinfections were not included.

### Exposure

Air pollutant concentrations for PM_10_, PM_2.5_, NO_2_ and NO_x_ were extracted from the Department of Environment, Food and Rural Affairs (DEFRA). DEFRA’s pollution climate mapping provides annual mean concentration at 1×1 kilometres (km) resolution, linked to a grid code with northings and eastings. ^21^ Long term exposure was defined as a five year average (2015-19) with separate sensitivity analysis using one year (2019) as the exposure. Annual mean concentrations up to 2019 were used as air pollution concentration changed due to COVID-19 related lockdown/restrictions in 2020. The northings and eastings were converted to longitude and latitude and then matched to participants’ postcodes, using longitude and latitude coordinates from the Office for National Statistics (ONS). The participants’ residential postcode were matched to pollutant concentration for each year by nearest latitude and longitude coordinates for each year from 2015-19, after which a five year average was taken. To check if the effect estimates were affected by the crude spatial resolution of the DEFRA air pollution source, the ELAPSE (Effect of Low-level Air Pollution: A Study in Europe) modelled data was used as part of the sensitivity analysis for PM_2.5_ and NO_2_. This data also contained estimates for BC, a component of PM_2.5_ and a specific marker for traffic-related air pollution in Europe, so was included in the analysis. The ELAPSE high resolution data (100×100 m) was created using a novel combination of land use regression and dispersion models, routine monitoring data and satellite observations.^22^

### Covariates

Potential confounders were identified using directed acyclic graphs (DAG; supplementary information Figure 1) using DAGitty, to provide minimally adjusted unbiased estimates of the total and direct effect.^23^ For estimating total effect two adjustment sets were used;

#### Adjustment set 1

Age & Sex, Socioeconomic status (i.e. household income), Ethnicity, Migrant Status, Occupation, Population Density (i.e. urban vs rural)

#### Adjustment set 2

Age & Sex, Socioeconomic status (i.e. household income), Geographical Region, Migrant Status, Occupation, Population Density (i.e. urban vs rural)

To estimate direct effect, comorbidities were added to each adjustment set as a binary variable. Comorbidities were defined as an individual having at least one of the following conditions; respiratory conditions, cardiovascular disease, diabetes, any cancer and obesity. Collapsing of data collection variables into categories, e.g., respiratory and cardiovascular diseases, followed the same approach as previous Virus Watch analyses.^24^ Socioeconomic status was defined by the reported combined household income, which was then categorised into quintiles. Population density was defined as urban or rural settings based on the ONS 2011 Rural-Urban Classification for Output Areas in England, defining urban areas as ‘connected built up areas identified by Ordnance Survey mapping that have resident populations above 10,000 people’.^25^ Occupation was collapsed into the following categories based on UK Standard Occupational Classification 2020 codes: administrative and secretarial occupations; healthcare occupations; indoor trade, process and plant occupations; leisure and personal service occupations; managers, directors, and senior officials; outdoor trade occupations; sales and customer service occupations; social care and community protective services; teaching education and childcare occupations; transport and mobile machine operatives; and other professional and associate occupations (professional and associate professional occupations excluding healthcare, teaching, and social care/community protective services).^26^

### Statistical and sensitivity analysis

Summary data was reported as means with standard deviations (SD) and proportions. Statistical tests to compare covariates between positive and negative cases were Welch’s two sample t-test for continuous data and Pearson’s Chi Squared test for categorical data. Pearson’s correlation coefficient was used to assess collinearity for annual mean air pollutant concentrations. We used a modified Poisson regression model with robust estimates, clustered by household, which is an established method for assessing relative risk using cohort data with a binary outcome.^27^ Regression analysis with minimal adjustments, adjustments for total and direct effects were carried out using a five year average (2015-2019) from DEFRA’s modelled air pollution and ELAPSE modelled air pollution data. Sensitivity analyses for estimating direct effect were carried out based on three assumptions; time frame of measurement, source of modelled air pollution estimates and inclusion of children into analysis. ‘Long-term exposure’ is not unanimously defined, so a five year average (2015-2019) and one year average (2019) were used. Different techniques are used to model air pollutant concentration for high resolution maps, including variation in mixture of observational sources used (e.g., ground-level or satellite) to ‘improve’ accuracy of estimates. Finally, compared to adults, children receive higher doses of air pollution due to their faster respiratory rate and higher intake per kilogram of bodyweight.^28,29^ Although age was adjusted for in our regression analysis, an analysis of just adults was also carried out. Regression analysis included both single-pollutant and bi-pollutant models. Only PM_2.5_ and NO_2_ were included in the bi-pollutant model as interaction terms. These two pollutants were selected as they are composites of PM_10_ and NO_x_ (respectively), were available in ELAPSE data for sensitivity analysis and replicates other analyses allowing for comparison of outcomes.^30^ Correlation coefficient was 0.76 and 0.69 between these two pollutants in DEFRA and ELAPSE data, respectively (supplementary information Figure 2 and 3).

### Data management

The modified Poisson regression analysis was carried out in Stata V17.0 using the mepoisson function. All other data management, including wrangling, analysis and visualisation was carried out in R 4.1.2 using the following packages; *tidyverse, ggplot2, psych, lubridate, gtsummary* and *hutilscpp*.

## Results

Of the 58,627 participants, we excluded 414 who had a positive SARS-CoV-2 test before the start of the second wave. 4,530 (mean age 41.2 (22.1), 1.1% SARS-CoV-2 positive) participants did not have a postcode that matched the ONS records, so were also excluded (Figure 1).

**Figure 1.**
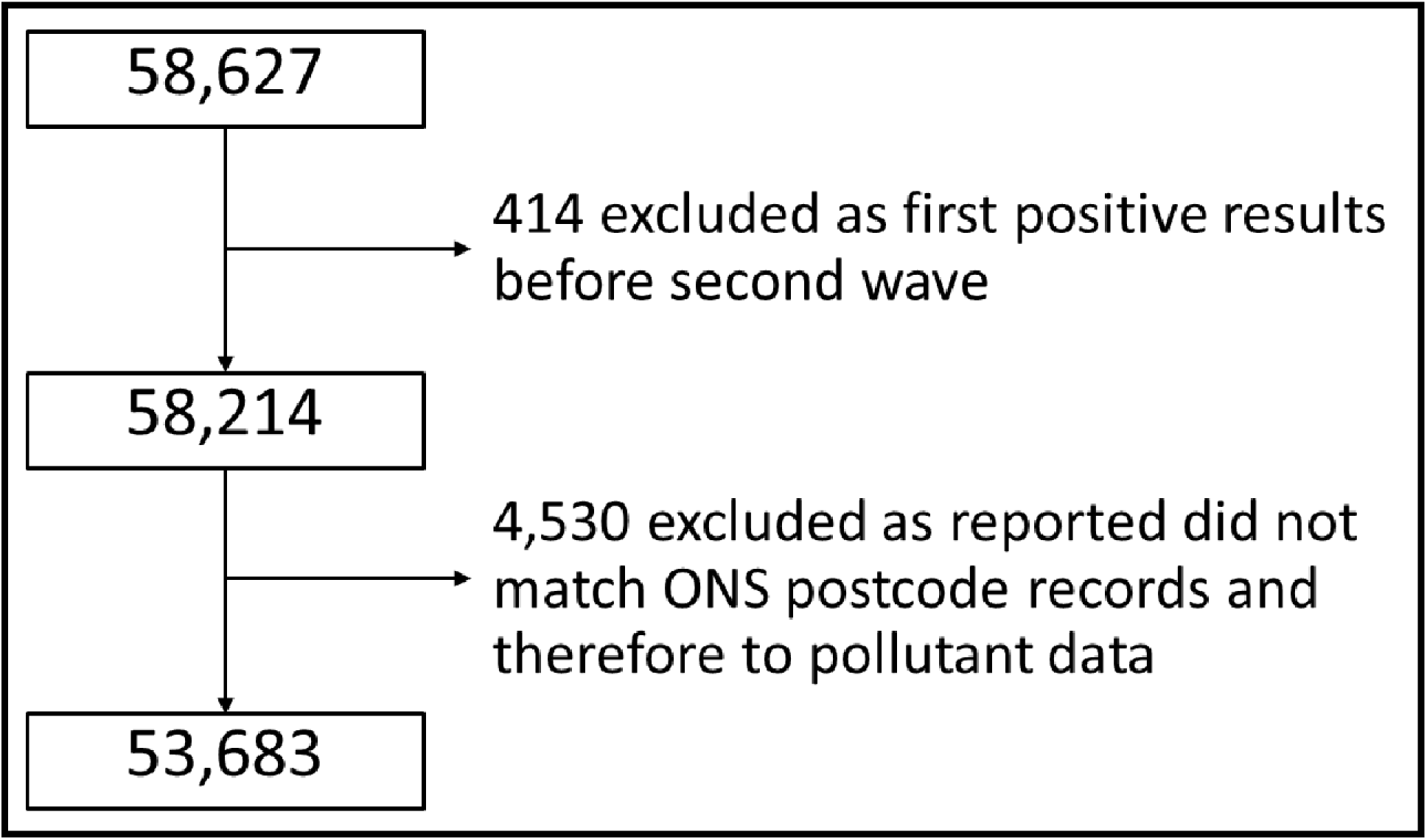
Flowchart of exclusion process of Virus Watch participants for analysis.

Therefore 53,683 participants were included in the analysis, with a mean age of 48.5 (21.77) and 53% (28,189) were female. 6.22% (3,341) of participants had a positive SARS-CoV-2 test during the second wave. Compared to those without SARS-CoV-2 infection, the positive group had a lower mean age, higher proportion of non-White British individuals, higher proportion of non-UK born participants and higher proportion of individuals in the lower quintiles of combined household income (p < 0.001, Table 1). There were also differences between the two groups for household overcrowding, regional location, urban vs rural setting, occupation as well as respiratory conditions, ischaemic heart disease, diabetes, cancer and obesity (supplementary information Table 1).

**Table 1.** Demographic information for all participants, then grouped by positive and negative SARS-CoV-2 groups, with comparison of covariates between groups.

| Characteristic | Overall, N = 53,683 | Negative, N = 50,342 | Positive, N = 3,341 | p-value<sup>1</sup> |
| --- | --- | --- | --- | --- |
| Age |  |  |  | <0.001 |
| Mean (SD) | 48.50 (21.77) | 48.66 (21.94) | 46.11 (18.93) |  |
| Age Group, n (%) |  |  |  | <0.001 |
| 0-17 | 7,476 (14%) | 7,146 (14%) | 330 (9.9%) |  |
| 35-49 | 9,320 (17%) | 8,530 (17%) | 790 (24%) |  |
| 18-34 | 6,540 (12%) | 5,944 (12%) | 596 (18%) |  |
| 50-64 | 14,796 (28%) | 13,798 (27%) | 998 (30%) |  |
| 65-79 | 14,531 (27%) | 13,937 (28%) | 594 (18%) |  |
| 80+ | 1,020 (1.9%) | 987 (2.0%) | 33 (1.0%) |  |
| Missing | 0 (0%) | 0 (0%) | 0 (0%) |  |
| Sex, n (%) |  |  |  | <0.001 |
| Male | 23,040 (43%) | 21,580 (43%) | 1,460 (44%) |  |
| Female | 28,189 (53%) | 26,319 (52%) | 1,870 (56%) |  |
| Intersex/Missing | 2,454 (4.6%) | 2,443 (4.9%) | 11 (0.3%) |  |
| Ethnicity, n (%) |  |  |  | <0.001 |
| Black | 479 (0.9%) | 436 (0.9%) | 43 (1.3%) |  |
| Mixed | 977 (1.8%) | 894 (1.8%) | 83 (2.5%) |  |
| Other Asian | 408 (0.8%) | 381 (0.8%) | 27 (0.8%) |  |
| Other Ethnicity | 285 (0.5%) | 242 (0.5%) | 43 (1.3%) |  |
| Prefer not to say | 189 (0.4%) | 175 (0.3%) | 14 (0.4%) |  |
| South Asian | 2,611 (4.9%) | 2,306 (4.6%) | 305 (9.1%) |  |
| White British | 39,620 (74%) | 37,306 (74%) | 2,314 (69%) |  |
| White Irish | 644 (1.2%) | 604 (1.2%) | 40 (1.2%) |  |
| White Other | 2,763 (5.1%) | 2,548 (5.1%) | 215 (6.4%) |  |
| Missing | 5,707 (11%) | 5,450 (11%) | 257 (7.7%) |  |
| UK Born, n (%) |  |  |  | <0.001 |
| UK Born | 48,498 (90%) | 45,650 (91%) | 2,848 (85%) |  |
| Not UK Born | 5,185 (9.7%) | 4,692 (9.3%) | 493 (15%) |  |
| Income band, n (%) |  |  |  | <0.001 |
| £0-24999 | 9,668 (18%) | 9,001 (18%) | 667 (20%) |  |
| £25000-£74999 | 18,777 (35%) | 17,449 (35%) | 1,328 (40%) |  |
| £75000-£124999 | 5,594 (10%) | 5,148 (10%) | 446 (13%) |  |
| £125000-£174999 | 1,193 (2.2%) | 1,106 (2.2%) | 87 (2.6%) |  |
| £175000 or more | 843 (1.6%) | 797 (1.6%) | 46 (1.4%) |  |
| Missing | 17,608 (33%) | 16,841 (33%) | 767 (23%) |  |
| Conditions associated with air pollution exposure, n (%) | 16,795 (30%) | 15,663 (30%) | 1,132 (33%) | <0.001 |

<sup>1</sup>Welch Two Sample t-test for continuous data; Pearson's Chi-squared test for categorical data

Taking a 5 year average of DEFRA’s annual mean air pollutant concentration (i.e., 2015-19), mean concentration for NO_2_, NO_x_, PM_10_ and PM_2.5_ was 15.53µg/m^3^ (6.81), 22.89µg/m^3^ (12.38), 14.74µg/m^3^ (2.5) and 12.97µg/m^3^ (1.57). Annual mean concentration for each air pollutant changed over time, the highest concentration for all air pollutants in 2016, but there was no linear trend (supplementary information Figure 4). The mean distance between location of pollutant concentration estimate and participant postcode was 350.34 (128.52) metres (m) for DEFRA’s 2015-19 data. Distances differed between DEFRA’s 2015-19 (350.34m, 95%CI 349;351) and 2019 (381.58m, 95%CI 380.38;382.78) data. Mean concentration in the positive group was higher for all air pollutants regardless of exposure window (i.e., DEFRA’s 2015-19 vs DEFRA’s 2019) and source (DEFRA vs ELAPSE), p < 0.001 (Table 2). Apart from NO_2_ (2015-19 and 2019) and NO_x_ (2019) when comparing combined household income quintiles of ‘£0-24999’ and ‘£75000-£124999’, mean concentration of each pollutant differed between quintiles with no linear trend using DEFRA data (supplementary information Figure 5).

**Table 2.** Mean pollutant concentration (µg m^-3^) for each.

| **Pollutant (Source)** | **Overall, N = 53,683**<br>Mean (SD) | **Negative, N = 50,342**<br>Mean (SD) | **Positive, N = 3,341**<br>Mean (SD) | **p-value<sup>1</sup>** |
| --- | --- | --- | --- | --- |
| NO<sub>2</sub> 2015-19 (DEFRA) | 15.53 (6.81) | 15.39 (6.74) | 17.64 (7.38) | <0.001 |
| NO<sub>2</sub> 2019 (DEFRA) | 14.86 (6.44) | 14.72 (6.38) | 16.88 (6.99) | <0.001 |
| NO<sub>2</sub> (ELAPSE) | 28.51 (8.01) | 28.34 (7.96) | 31.06 (8.23) | <0.001 |
| NO<sub>x</sub> 2015-19 (DEFRA) | 22.89 (12.38) | 22.65 (12.24) | 26.61 (13.79) | <0.001 |
| NO<sub>x</sub> 2019 (DEFRA) | 21.56 (11.74) | 21.33 (11.61) | 25.15 (13.13) | <0.001 |
| PM<sub>2.5</sub> 2015-19 (DEFRA) | 10.24 (1.59) | 10.22 (1.58) | 10.61 (1.63) | <0.001 |
| PM<sub>2.5</sub> 2019 (DEFRA) | 9.63 (1.74) | 9.60 (1.74) | 10.07 (1.78) | <0.001 |
| PM<sub>2.5</sub> (ELAPSE) | 12.97 (1.57) | 12.95 (1.57) | 13.25 (1.52) | <0.001 |
| PM<sub>10</sub> 2015-19 (DEFRA) | 14.74 (2.50) | 14.71 (2.48) | 15.28 (2.65) | <0.001 |
| PM<sub>10</sub> 2019 (DEFRA) | 14.99 (2.66) | 14.95 (2.65) | 15.61 (2.78) | <0.001 |
| BC (ELAPSE) | 1.33 (0.37) | 1.33 (0.37) | 1.44 (0.39) | <0.001 |

<sup>1</sup>Welch Two Sample t-test. PM<sub>10</sub> = particulate matter 10. PM<sub>2.5</sub> = particulate matter 2.5, NO<sub>2</sub> = nitrogen dioxide. NO<sub>x</sub> = oxides of nitrogen. BC = Black Carbon

Using DEFRA 2015-19 data, single pollutant modified Poisson regression analysis showed higher risk of SAR-CoV-2 infection with increased exposure to PM_2.5_, PM_10_, NO_2_ and NO_x_ when adjusted for age and sex (Figure 2). Estimates for total effect and direct effect were similar for PM_2.5_ (1.11, 95%CI 1.08;1.15) followed by PM_10_ (1.06, 95%CI 1.04;1.09), NO_2_ (1.04, 95%CI 1.04;1.05) and NO_x_ (1.02, 95%CI 1.02;1.02) per 1µg/m^3^ increment using adjustment set 1. Estimates between total and direct effects for adjustment set 2 were also similar: PM_2.5_ (1.25, 95%CI 1.17;1.33), PM_10_ (1.10, 95%CI 1.06;1.14), NO_2_ (1.04, 95%CI 1.02;1.04) and NO_x_ (1.01, 95%CI 1.01;1.02) per 1µg/m^3^ increment. Bi-pollutant modified Poisson regression analysis results showed an increase in risk per 1µg/m^3^ NO_2_ increment, apart from adjustment set 2 using 2019 DEFRA data. The higher risk of infection per 1µg/m^3^ increase in PM_2.5_ remained in adjustment set 2 for 2015-19 (1.22, 95%CI 1.09;1.36) and 2019 (1.19, 95%CI 1.08;1.30), but was not statistically significant in adjustment set 1 (Figure 3).

**Figure 2.**
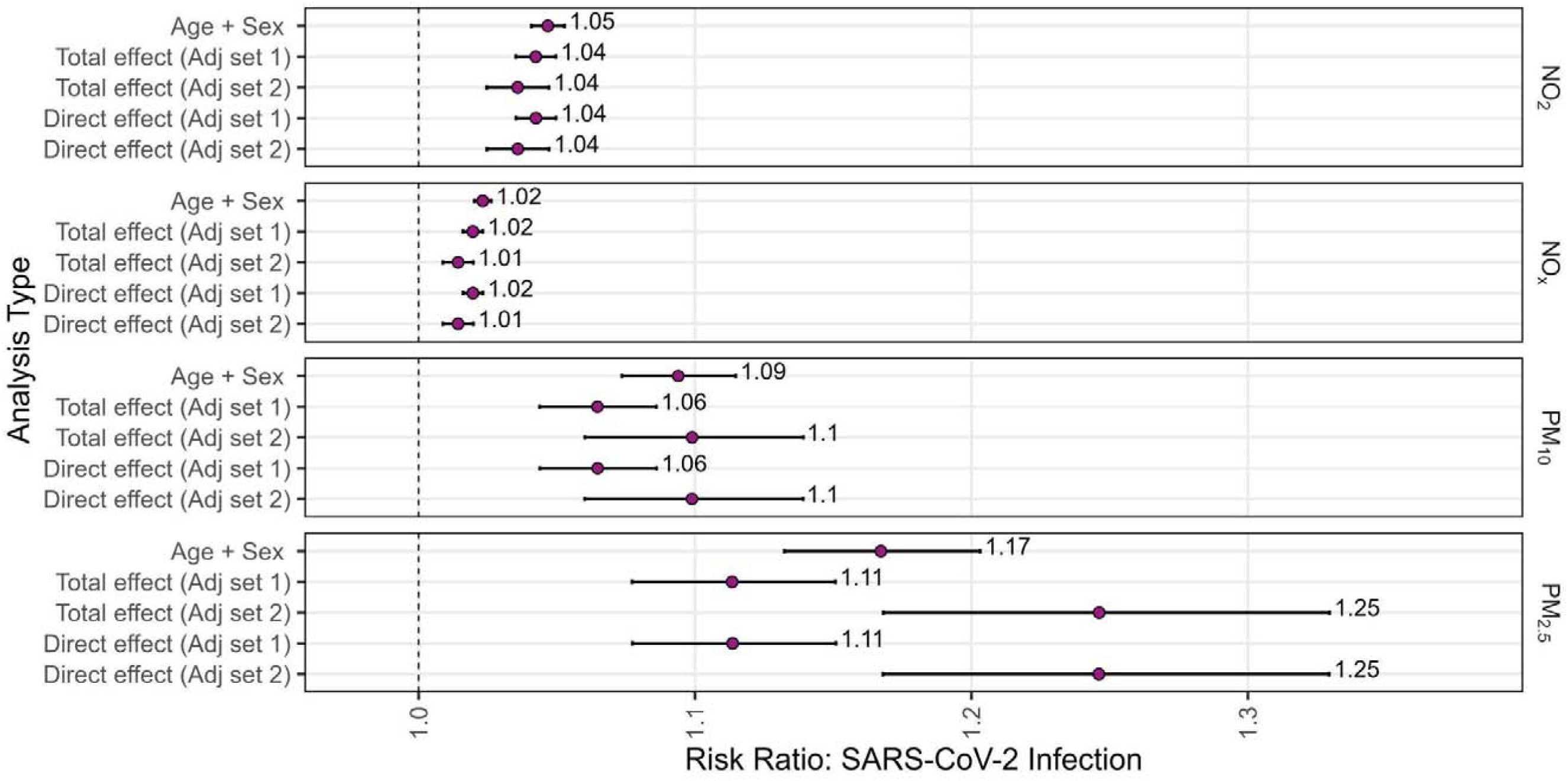
Risk of SARS-CoV-2 infection per one µg m^-3^ increment in single-pollutant model of long-term air pollution exposure (DEFRA, five year average 2015-2019), by pollutant.

**Figure 3.**
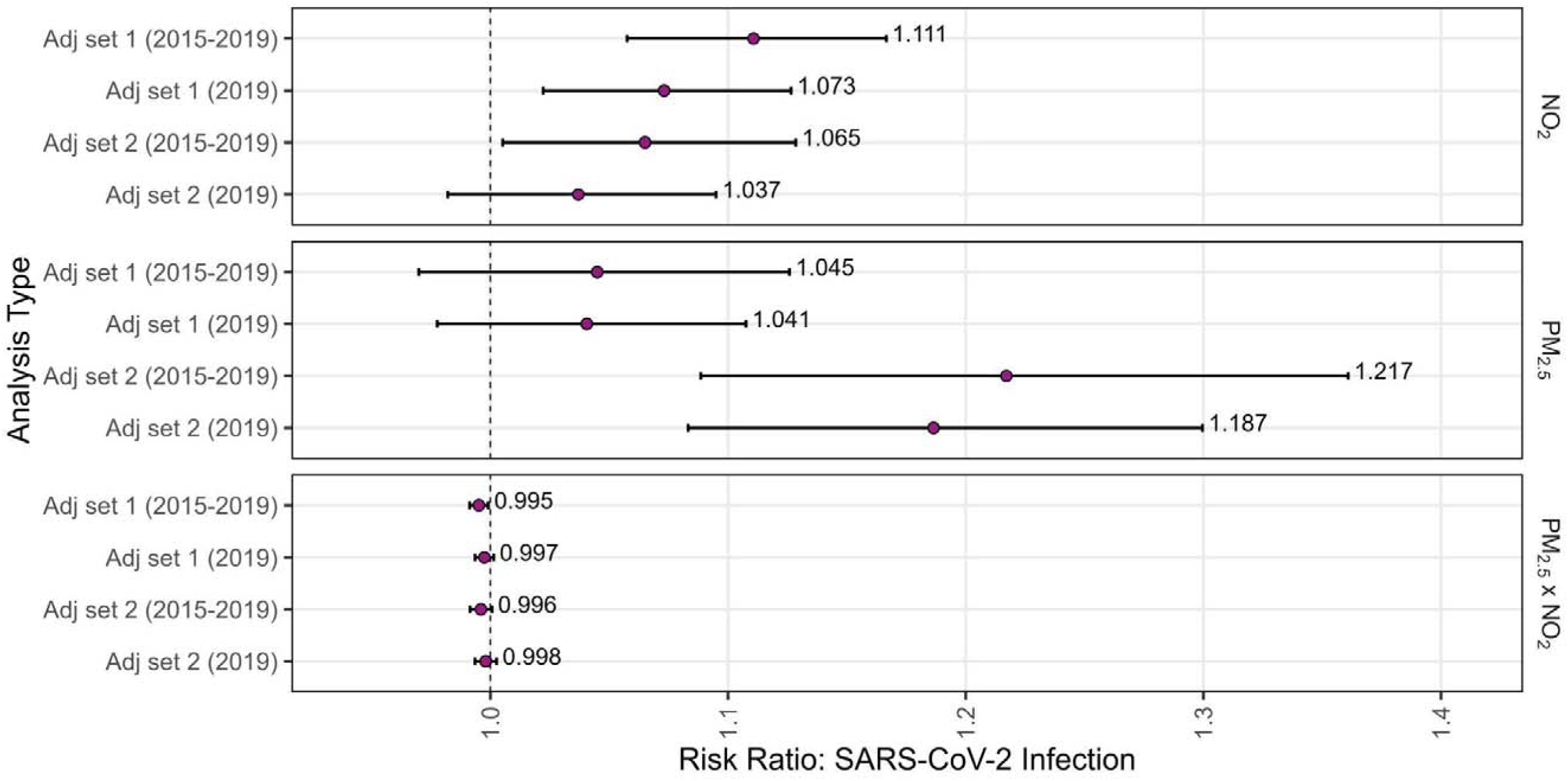
Risk of SARS-CoV-2 infection per one µg m^-3^ increment in bi-pollutant model (interaction terms) of long-term air pollution exposure (DEFRA), by pollutant, for all participants of the Virus Watch study.

Similar estimates of direct effect were seen between two exposure windows of five years (2015-19) and one year (2019), regardless of adjustment set 1 vs adjustment set 2 or all participants vs adults only (supplementary information Figure 7 and 8). Regression analysis with the ELAPSE data showed similar results to the DEFRA data for adjustment set 1, but risk of infection per 1µg/m^3^ increment PM_2.5_ was lower for adjustment set 2 (1.09, 95%CI 1.04;1.13) for both total and direct effect (Figure 4). As seen in estimates using modelled pollutant data from DEFRA, estimates of total effect and direct effect were similar for PM_2.5_, BC and NO_2_ from the ELAPSE data. Risk of SARS-CoV-2 from one 10^-5^m^-1^ increment of BC was 1.89 (95%CI 1.64;2.18), for total effect (adjustment set 1 and 2) and direct effect (adjustment set 1), but was 1.67 (95%CI 1.39;2.01) for direct effect (adjustment set 2). Apart from NO_2_ (adjustment set 1), the bi-pollutant regression analysis with the ELAPSE data showed no increased risk of SARS-CoV-2 infection (Figure 5).

**Figure 4.**
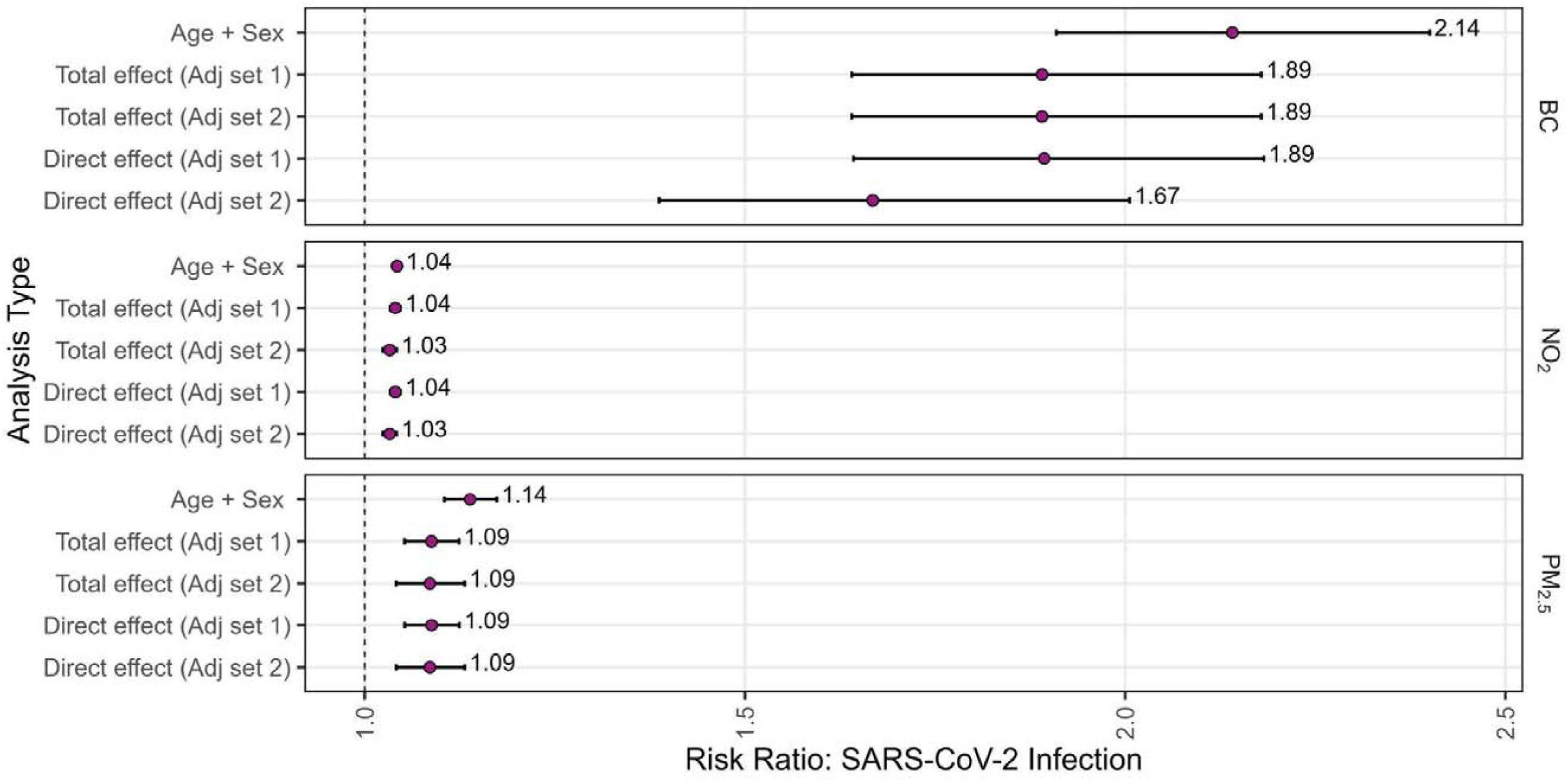
Risk of SARS-CoV-2 infection per one µg/m^3^ (except BC; 10^-5^m^-1^) increment in single-pollutant model of long-term air pollution exposure (ELAPSE), by pollutant for all participants of the Virus Watch study.

**Figure 5.**
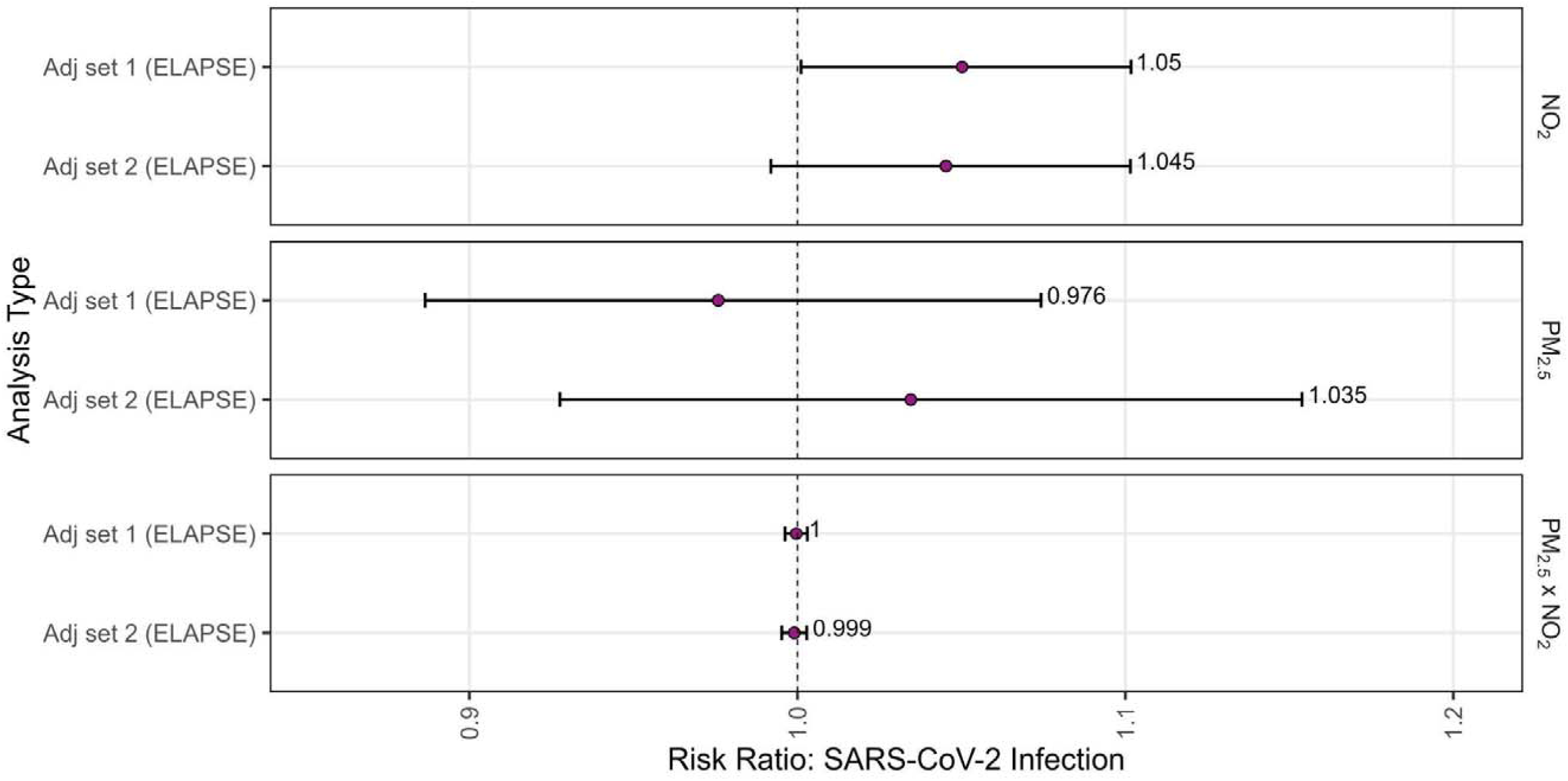
Risk of SARS-CoV-2 infection per one µg/m^3^ (except BC; 10^-5^m^-1^) increment in bi-pollutant model (interaction terms) of long-term air pollution exposure (ELAPSE), by pollutant, for all participants of the Virus Watch study.

## Discussion

This cohort study shows higher risk of SARS-CoV-2 infection with long term exposure to higher concentrations of BC, PM_2.5_, PM_10_, NO_2_ and NO_x_ when each pollutant is assessed in isolation, regardless of the air pollution data source used. This increased risk is evident when adjusted for total effect and direct effects, each giving consistent estimates. Regression analysis adjusted for individual covariates including age, sex, socioeconomic status, ethnicity, migrant status, occupation, geographical setting and underlying medical conditions. Sensitivity analysis, adjusting for direct effect, showed the increased risk of infection remained despite changes to potential confounders, time frame of exposure measurement or source of air pollution data.

There was minimal interaction between PM_2.5_ and NO_2_ in bi-pollutant model. Higher risk of infection was seen with increments of PM_2.5_ concentration in adjustment set 2 compared to adjustment set 1 (i.e., ethnicity was replaced with region as a covariate). This may be due to high spatial variability of PM_2.5_, so use of region in the adjustment set and data from lower resolution pollutant models (e.g., 1km x 1km, compared to 100m x 100m in ELAPSE) would impact the estimates.^31^

There have been several studies that have identified an increased risk of SARS-CoV-2 infection with increases in PM_2.5_, PM_10_ and NO_2_. Few however adjusted their estimates for confounding factors with individual level data and none have included BC. Evidence in the UK on long term air pollution exposure and risk of SARS-CoV-2 infection at the individual level derives from UK biobank.^32–35^ All of these analyses showed a positive association between long-term PM_2.5_ and NO_2_ exposure and risk of SARS-CoV-2 infection, but none have taken a causal approach. The only study to use individual-level data for socioeconomic status also only included ‘Pillar 1’ data from UK Health Security Agency (formerly Public Health England), a time of limited testing.^32^ During this time, testing was more likely in urban areas that have greater healthcare resources (i.e., easier access to testing) and higher air pollution levels, questioning potential overestimation of the positive association.^32,36^ The results of our analysis confirm this positive association, demonstrating a causal effect, during a time when testing was widely accessible in both urban and rural areas.

There was little difference in estimates when adjusting for total and direct effects (i.e., inclusion of comorbidities). Similar findings have been reported previously, corroborating direct pathways are relevant.^17,37,38^ The direct effects of air pollution on the immune system within lung tissue has been explored in human and in vitro studies. Diesel exhaust, which contains NO_x_, NO_2_ and BC, promotes eosinophilic activity and therefore allergic inflammation and may be associated with reduced virus clearance.^39^ Individuals with short term exposure to higher concentrations of NO_2_, had less effective inactivation of influenza virus on analysis of their bronchoalveolar lavage 3.5 hours later.^40^ PM_2.5_ containing redox-active transition metals, quinones and secondary organic aerosols can increase reactive oxygen species (ROS) in the epithelial lining fluid of the respiratory tract, similar to levels characteristic of respiratory diseases.^41^ This oxidative stress may also decrease phagocytic function of macrophages impairing immune functions of the lungs.^42^ PM_2.5_ may also impair immune function through inhibition of airway antimicrobial peptide expression, specifically salivary agglutinin and surfactant protein D, and facilitate pathogen adhesion to airway epithelial cells, resulting in host pathogen infection.^43^ Mechanisms specific to SARS-CoV-2 have also been proposed, associating long-term PM_2.5_ exposure with increased alveolar expression of ACE2, increasing the chance of viral receptor binding and entry into host cells.^6,9,10,44^

Our findings require cautious interpretation in causal terms, as there remains a risk of residual confounding within our estimates. Analysis was focussed on long-term exposure as we could not account for exposure from travel patterns, duration and location of occupational, educational or leisure activities. We made an important assumption - which is made by most analyses on the association of long-term air pollution exposure and health outcomes - that individual exposure was predominantly at residential addresses and these addresses were permanent (i.e., at least one year prior to registration). This isn’t entirely inappropriate however, as COVID restrictions took place during this study period, so many people were working from home. Additionally, concentration of air pollution decreased during 2020 as a result of COVID restrictions, so estimates from 2019 or earlier are more reflective of a ‘business as usual’ environment. Results of the bi-pollutant model should be interpreted cautiously as there is no unanimous agreement on bi-pollutant and multi-pollutant regression analyses, due to conflicting opinions on the importance of interactions and collinearity.^45–49^ We included pollutants that were chemically different to one another and selected those with the lowest correlation coefficients, although still high at around 0.7, as well as accounted for potential interaction. Although we adjusted for covariates such as migrant status and ethnicity in our regression analysis, it must be acknowledged that the cohort is predominantly White-British UK-born participants and likely more affluent than the general population. Duration and severity of co-morbidities were also not accounted for, but any diagnoses after 30th April 2021 were not included in the analysis.

There are several strengths to this analysis. Our cohort consists of 53,683 individuals and we only included cases that were confirmed either by serology or PCR. We limited the analysis of this cohort study to the second wave of the COVID-19 pandemic to minimise confounding of testing and vaccine accessibility. Our cohort consists of participants from a wide range of ethnicities, occupation and geographical settings (i.e., England and Wales, urban and rural), therefore more generalisable than other city or workforce focused analyses. All covariates were based on individual or household level data, minimising residual confounding, and regression analysis accounted for household clustering. Our sensitivity analyses demonstrated a higher risk of SARS-CoV-2 infection associated with four air pollutants, regardless of exposure window, cohort make up (i.e., adults only or all participants) and sources of modelled pollutant data. In addition to the novelty of multiple sensitivity analyses to this causal approach, we also included BC into our analysis, a traffic-related pollutant that has been overlooked despite its known implications on health outcomes.^50^

## Conclusion

There is a higher risk of SARS-CoV-2 infection as a result of long term exposure to higher concentrations of BC, PM_2.5_, PM_10_, NO_2_ and NO_x_. Estimates for total and direct effect were similar, suggesting direct pathways may play a key role to risk of infection. Effect estimates were similar despite variations in exposure window, covariate adjustment and sources of modelled air pollution data for PM_2.5_, PM_10_, NO_2_ and NO_x_. This study contributes evidence to the causal effects of air pollution on risk of SARS-CoV-2 infection with additional novelty of sensitivity analyses and inclusion of BC.

## Ethics approval

This study involves human participants and Virus Watch was approved by the Hampstead NHS Health Research Authority Ethics Committee (20/HRA/2320) and conformed to the ethical standards set out in the Declaration of Helsinki. Participants gave informed consent to participate in the study before taking part.

## Data availability

We aim to share aggregate data from this project on our website and via a “Findings so far” section on our website - https://ucl-virus-watch.net/. We will also be sharing individual record level data on a research data sharing service such as the Office of National Statistics Secure Research Service. In sharing the data we will work within the principles set out in the UKRI Guidance on best practice in the management of research data. Access to use of the data whilst research is being conducted will be managed by the Chief Investigators (ACH and RWA) in accordance with the principles set out in the UKRI guidance on best practice in the management of research data. We will put analysis code on publicly available repositories to enable their reuse.

## Contributors

Conceptualization (AMDN, RA, AH), Data curation (AMDN, SB, VN, AY, EF, CG), Formal Analysis (AMDN), Funding acquisition (RA, AH), Methodology (AMDN, YB, SB, HK, RA, AH), Project administration (JK), Software (VN, CG), Supervision (HK, RA, AH), Validation (SB), Visualization (AMDN), Writing − original draft (AMDN), Writing − reviewing and editing (all)

