## Supplementary files for "Assessing the causal effect of air pollution on risk of SARS-CoV-2 infection"

### Supplementary Information

Figure 1. Directed Acylic Graph with air pollution as the exposure of interest and SARS-CoV-2 infection as the outcome.

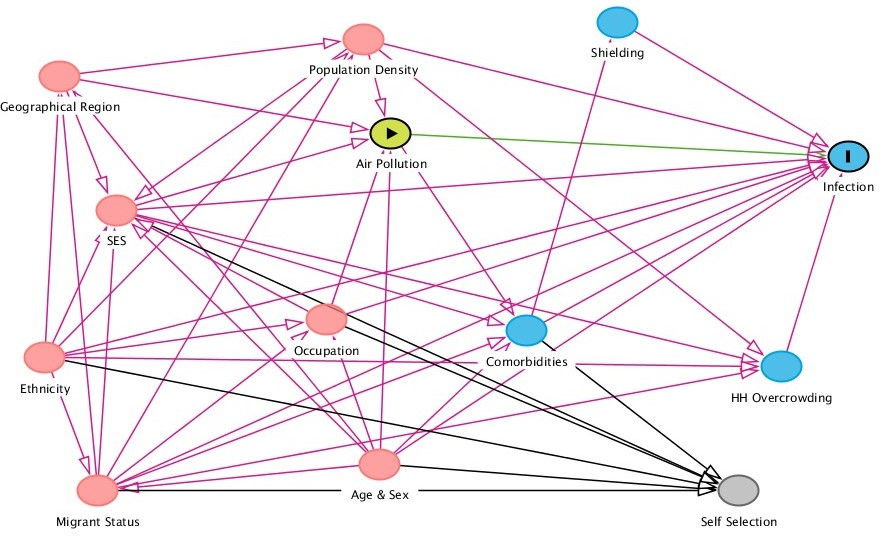

Figure 2. Correlation plots and coefficient between each pollutant (DEFRA, a five year average between 2015-19), with red line is local regression line (locally estimated scatterplot smoothing).

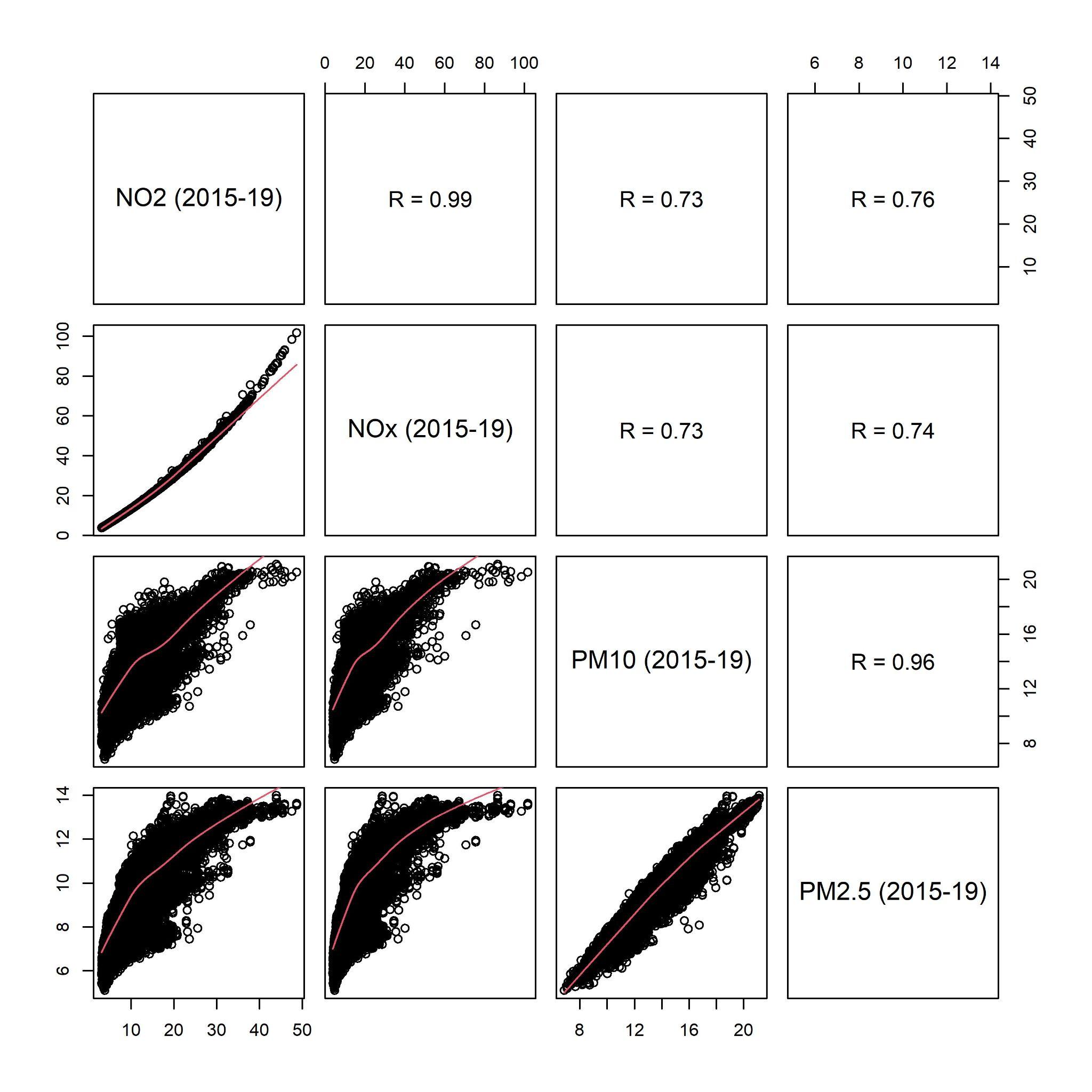

Figure 3. Correlation plots and coefficient between each pollutant (DEFRA 2019), with red line is local regression line (locally estimated scatterplot smoothing).

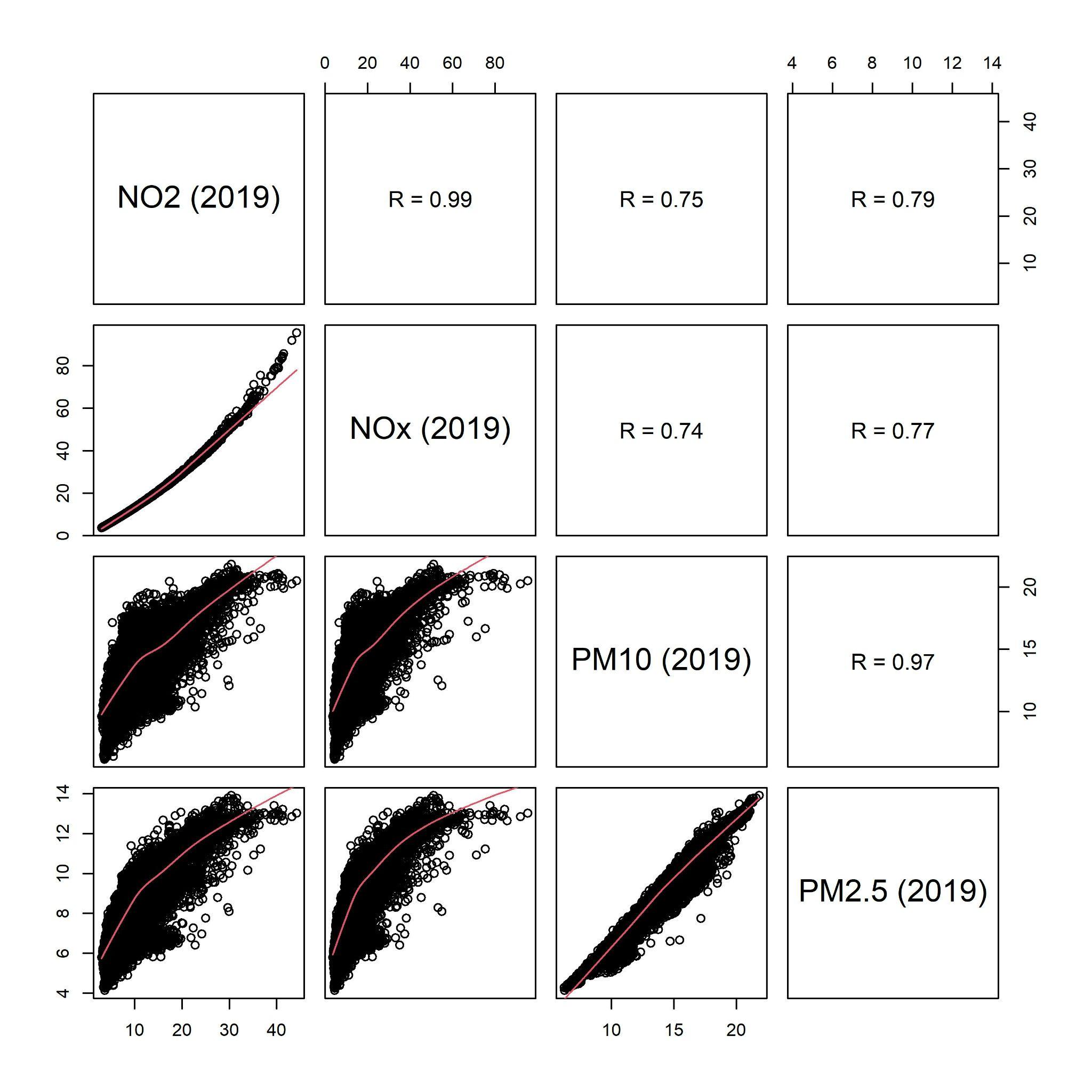

Figure 4: Correlation plots and coefficient between each pollutant (ELAPSE 2010), with red line is local regression line (locally estimated scatterplot smoothing).

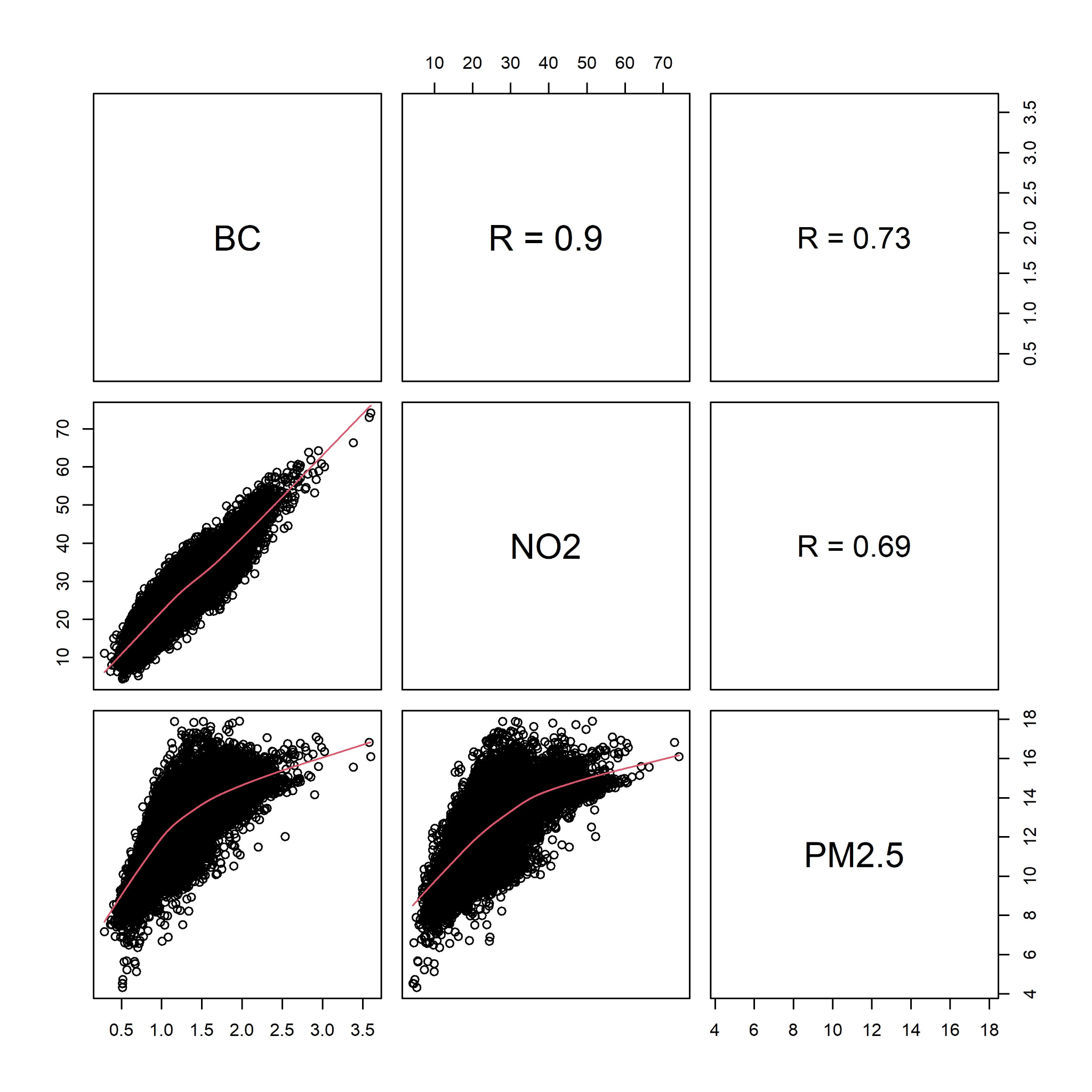

Figure 5. Mean air pollutant concentration linked to a participant of the Virus Watch study per pollutant per year. Annual mean concentration was extracted from DEFRA and linked to each participant by latitude and longitude postcode and estimate location.

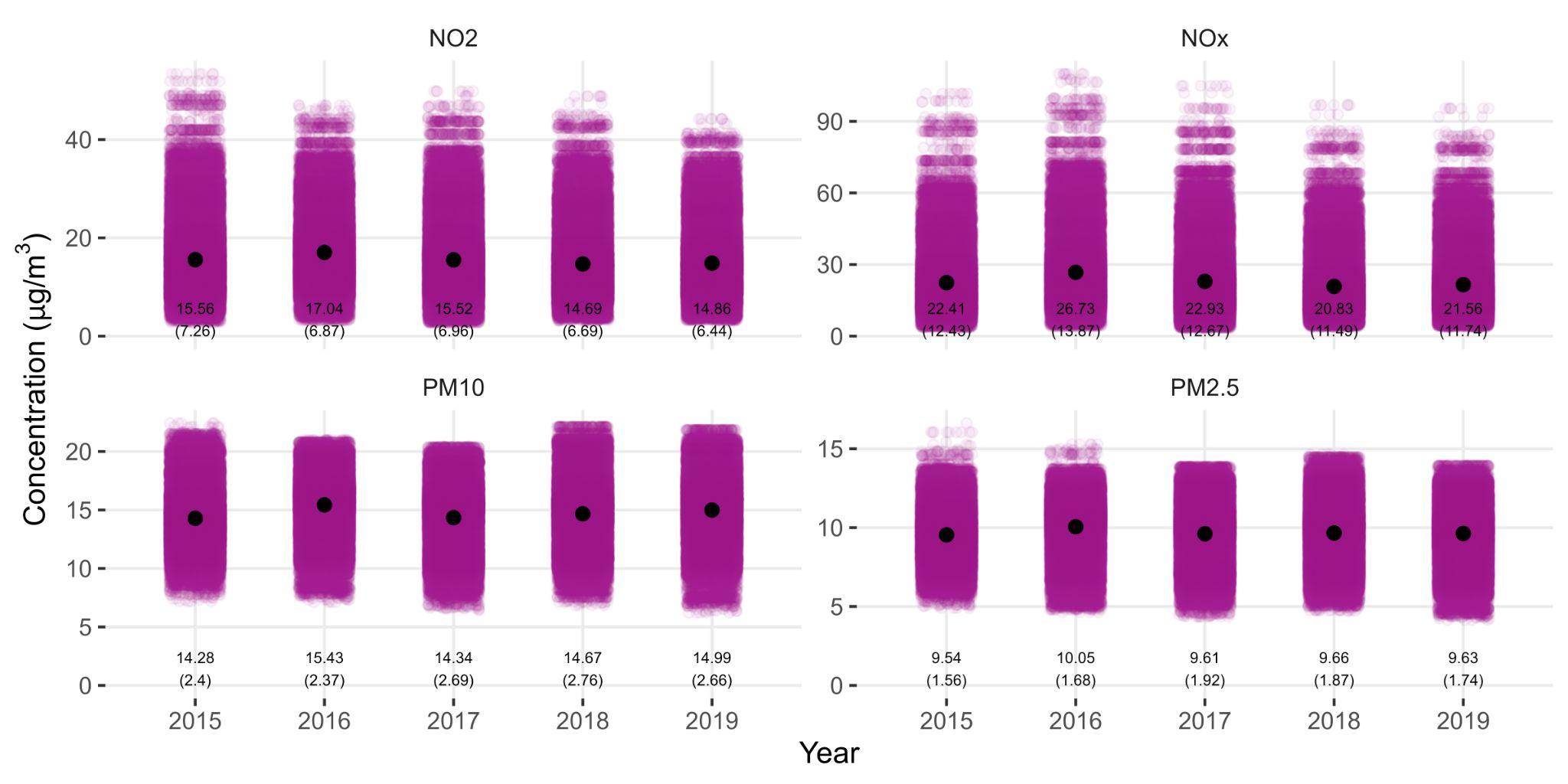

Figure 6. Mean air pollutant (black dots) concentration for each quintile of household income, with bars representing the 95% confidence intervals. There are significant differences between each group, but it does not follow a linear trend (i.e., as income reduces, air pollutant concentration increases).

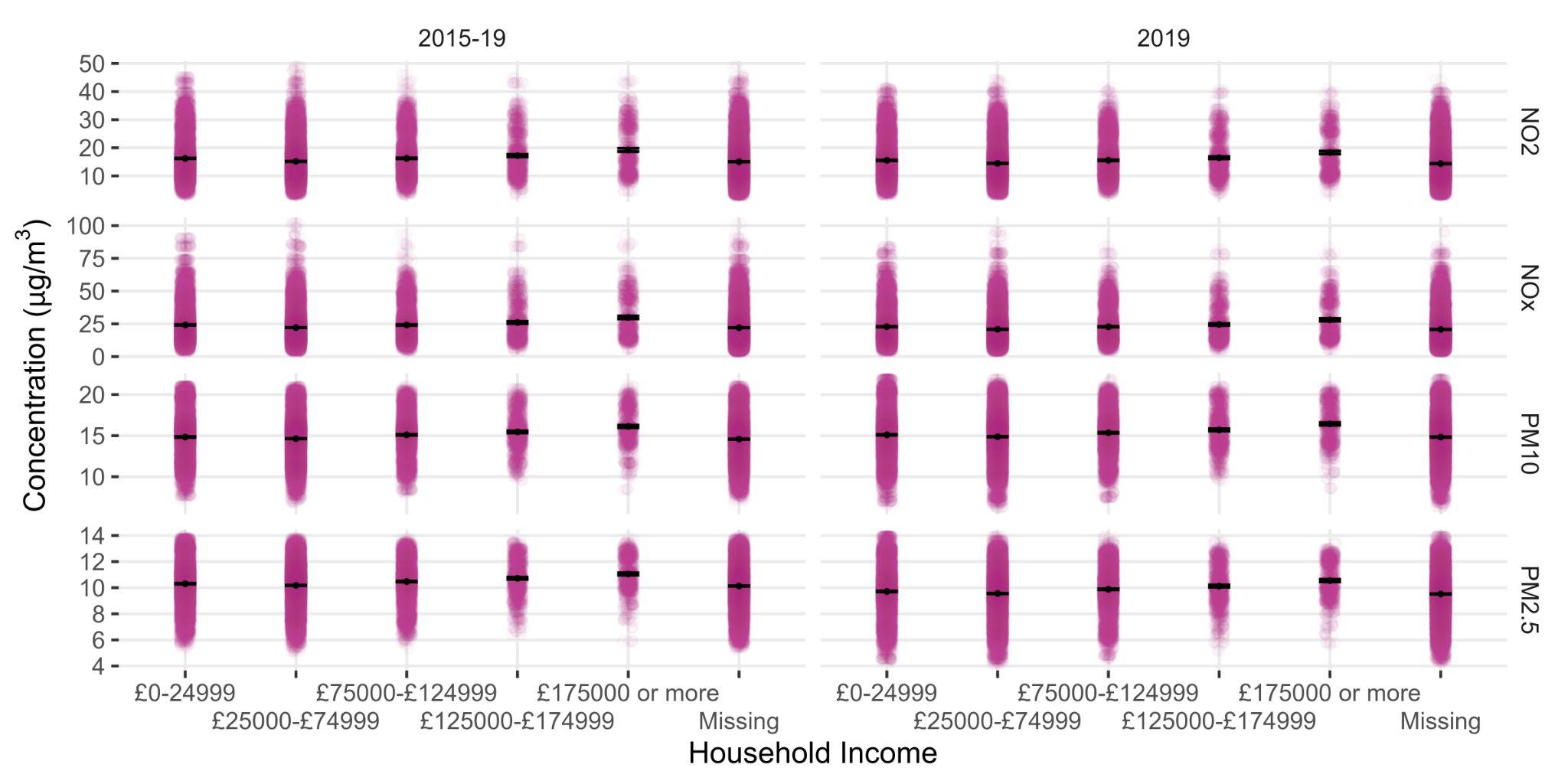

Figure 7. Risk of SARS-CoV-2 infection per one µg/m^3^ increment in single-pollutant model of long-term air pollution exposure (DEFRA), by pollutant, for all participants of the Virus Watch study.

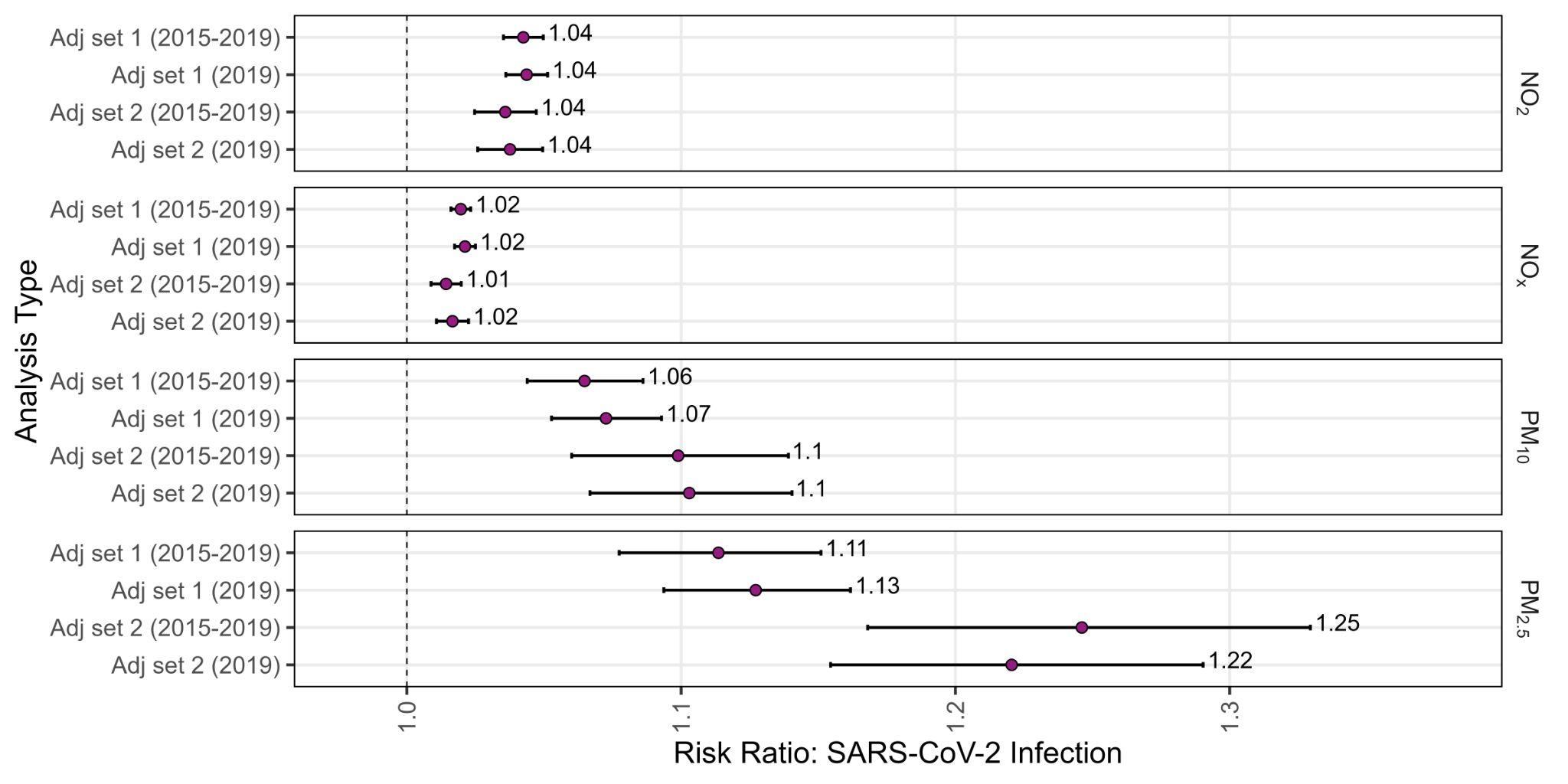

Figure 8. Risk of SARS-CoV-2 infection per one µg m^-3^ increment in single-pollutant model of long-term air pollution exposure (DEFRA), by pollutant, for only adults (⩾ 18 years old) participating in the Virus Watch study.

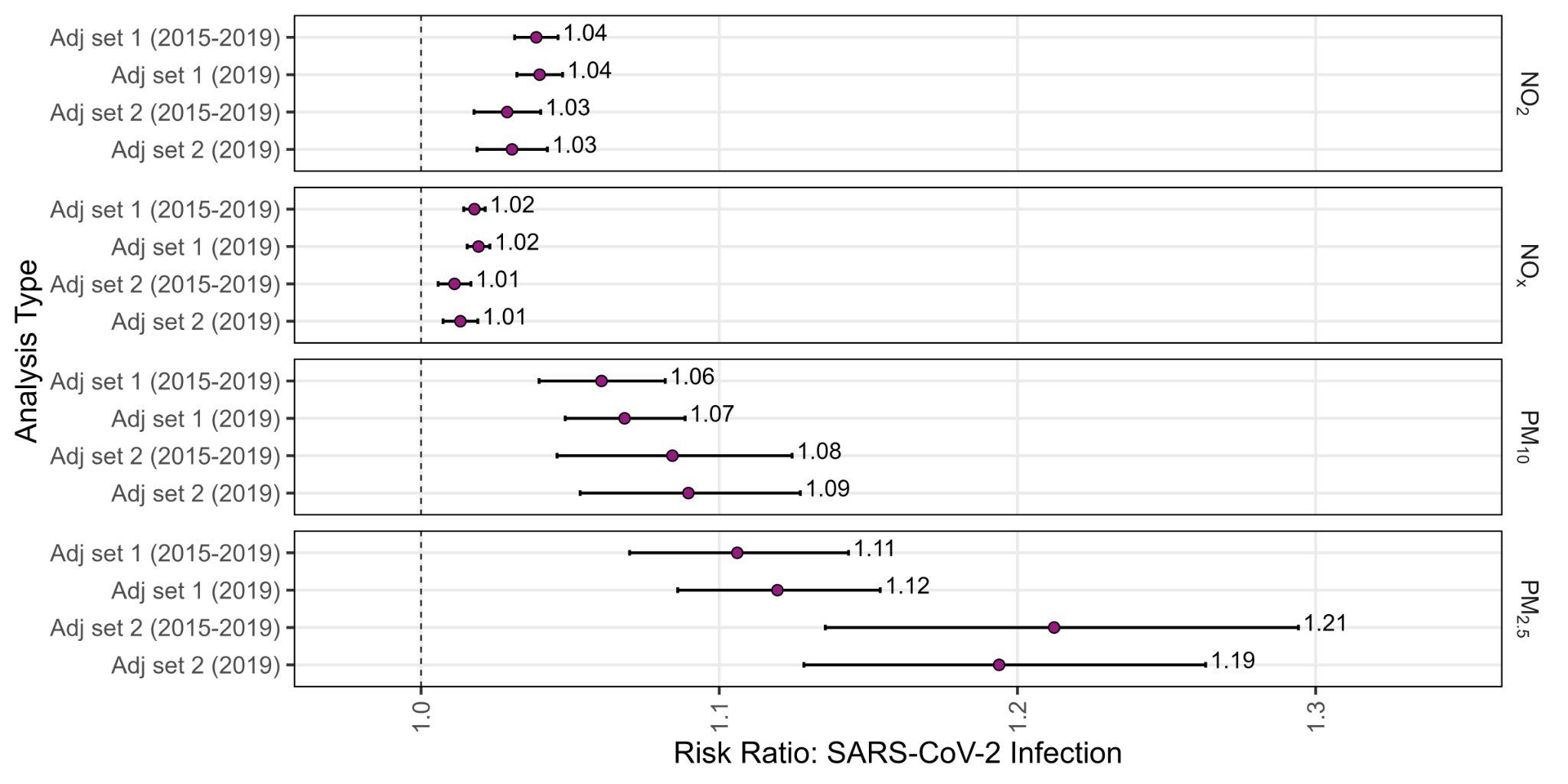

Table 1. Information for all participants, then grouped by positive and negative SARS-CoV-2 groups, with comparison of covariates between groups.

| **Characteristic** | **Overall**, N = 53,683 | **Negative**, N = 50,342 | **Positive**, N = 3,341 | **p-value**^1^ |
| --- | --- | --- | --- | --- |
| Overcrowding, n (%) |  |  |  | <0·001 |
| Balanced | 2,097 (3·9%) | 1,908 (3·8%) | 189 (5·7%) |  |
| Overcrowded | 952 (1·8%) | 856 (1·7%) | 96 (2·9%) |  |
| Under Occupied | 22,718 (42%) | 21,350 (42%) | 1,368 (41%) |  |
| Missing | 27,916 (52%) | 26,228 (52%) | 1,688 (51%) |  |
| Region, n (%) |  |  |  | <0·001 |
| East Midlands | 4,905 (9·1%) | 4,634 (9·2%) | 271 (8·1%) |  |
| East of England | 10,490 (20%) | 9,905 (20%) | 585 (18%) |  |
| London | 8,984 (17%) | 8,106 (16%) | 878 (26%) |  |
| North East | 2,512 (4·7%) | 2,370 (4·7%) | 142 (4·3%) |  |
| North West | 5,532 (10%) | 5,161 (10%) | 371 (11%) |  |
| South East | 9,784 (18%) | 9,276 (18%) | 508 (15%) |  |
| South West | 3,933 (7·3%) | 3,785 (7·5%) | 148 (4·4%) |  |
| Wales | 1,514 (2·8%) | 1,460 (2·9%) | 54 (1·6%) |  |
| West Midlands | 3,000 (5·6%) | 2,813 (5·6%) | 187 (5·6%) |  |
| Yorkshire and The Humber | 2,997 (5·6%) | 2,802 (5·6%) | 195 (5·8%) |  |
| Missing | 32 (<0·1%) | 30 (<0·1%) | <5 (<0·1%) |  |
| Urban/Rural, n (%) |  |  |  | <0·001 |
| Rural | 11,768 (22%) | 11,240 (22%) | 528 (16%) |  |
| Urban | 41,883 (78%) | 39,072 (78%) | 2,811 (84%) |  |
| Missing | 32 (<0·1%) | 30 (<0·1%) | <5 (<0·1%) |  |
| Occupation, n (%) |  |  |  | <0·001 |
| Other professional & associate | 6,406 (12%) | 5,956 (12%) | 450 (13%) |  |
| Administrative & Secretarial | 2,494 (4·6%) | 2,312 (4·6%) | 182 (5·4%) |  |
| Healthcare | 1,585 (3·0%) | 1,411 (2·8%) | 174 (5·2%) |  |
| Indoor Trades, Process & Plant | 1,371 (2·6%) | 1,241 (2·5%) | 130 (3·9%) |  |
| Leisure & Personal Service | 986 (1·8%) | 877 (1·7%) | 109 (3·3%) |  |
| Managers, Directors & Senior Officials | 1,622 (3·0%) | 1,511 (3·0%) | 111 (3·3%) |  |
| Outdoor Trades | 465 (0·9%) | 437 (0·9%) | 28 (0·8%) |  |
| Sales & Customer Service | 1,033 (1·9%) | 935 (1·9%) | 98 (2·9%) |  |
| Social Care & Community Protective Services | 1,081 (2·0%) | 983 (2·0%) | 98 (2·9%) |  |
| Teaching, Education & Childcare | 2,253 (4·2%) | 2,028 (4·0%) | 225 (6·7%) |  |
| Transport & Mobile Machine | 470 (0·9%) | 430 (0·9%) | 40 (1·2%) |  |
| Missing | 33,917 (63%) | 32,221 (64%) | 1,696 (51%) |  |
| ^+^Shielding, n (%) |  |  |  | <0·001 |
| No | 17,265 (32%) | 16,064 (32%) | 1,201 (36%) |  |
| Yes | 1,591 (3·0%) | 1,506 (3·0%) | 85 (2·5%) |  |
| Missing | 34,827 (65%) | 32,772 (65%) | 2,055 (62%) |  |
| Respiratory condition, n (%) |  |  |  | <0·001 |
| No | 35,080 (65%) | 32,691 (65%) | 2,389 (72%) |  |
| Yes | 7,007 (13%) | 6,548 (13%) | 459 (14%) |  |
| Missing | 11,596 (22%) | 11,103 (22%) | 493 (15%) |  |
| Ischaemic Heart Disease, n (%) |  |  |  | <0·001 |
| No | 40,500 (75%) | 37,740 (75%) | 2,760 (83%) |  |
| Yes | 1,587 (3·0%) | 1,499 (3·0%) | 88 (2·6%) |  |
| Missing | 11,596 (22%) | 11,103 (22%) | 493 (15%) |  |
| Diabetes, n (%) |  |  |  | <0·001 |
| No | 39,716 (74%) | 37,034 (74%) | 2,682 (80%) |  |
| Yes | 2,371 (4·4%) | 2,205 (4·4%) | 166 (5·0%) |  |
| Missing | 11,596 (22%) | 11,103 (22%) | 493 (15%) |  |
| Obese, n (%) |  |  |  | <0·001 |
| No | 24,944 (46%) | 23,270 (46%) | 1,674 (50%) |  |
| Yes | 7,365 (14%) | 6,790 (13%) | 575 (17%) |  |
| Missing | 21,374 (40%) | 20,282 (40%) | 1,092 (33%) |  |
| Cancer, n (%) |  |  |  | 0·002 |
| No | 27,657 (50%) | 25,877 (50%) | 1,780 (52%) |  |
| Yes | 2,303 (4·2%) | 2,193 (4·2%) | 110 (3·2%) |  |
| Missing | 25,458 (46%) | 23,928 (46%) | 1,530 (45%) |  |

^+^Defined being advised not leave their homes and minimising all face-to-face contact
